## Supplementary material for "Mitigating the Opioid Epidemic: The Role of Cannabinoids in Chronic Pain Management—A Systematic Review and Meta-Analysis of Clinical Evidence and Mechanisms": Suplemmental Materials

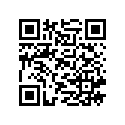

Author: Borges, Julian Yin Vieira M.D

Board Certified Endocrinologist, Board Certified in Medical Nutrition

Research Physician <https://orcid.org/0009-0001-9929-3135>

Supplemental Materials

Search Strategy

To ensure a comprehensive and systematic search for articles related to the efficacy of medicinal cannabis in chronic pain management and its comparison with opioids, the following strategy can be employed:

Databases

- PubMed
- Embase
- Cochrane Library
- PsycINFO
- Web of Science

Keywords and Search Terms

1. Pain Management:
   - Chronic pain
   - Pain relief
   - Pain control
   - Pain therapy
2. Cannabinoids:
   - Medical cannabis
   - Cannabidiol (CBD)
   - Tetrahydrocannabinol (THC)
   - Nabiximols
3. Opioids:
   - Opioid analgesics
   - Opioid epidemic
   - Opioid crisis
   - Opioid therapy
4. Efficacy and Safety:
   - Efficacy
   - Safety
   - Adverse effects
   - Comparative effectiveness
5. Study Design:
   - Randomized controlled trials
   - Systematic reviews
   - Meta-analyses
   - Observational studies

Search Strings

1. PubMed:

mathematica

Copy code

(chronic pain[MeSH Terms] OR chronic pain[Text Word]) AND (cannabinoids[MeSH Terms] OR medical cannabis[Text Word] OR cannabidiol[Text Word] OR tetrahydrocannabinol[Text Word] OR nabiximols[Text Word]) AND (opioid[MeSH Terms] OR opioid[Text Word]) AND (efficacy[Text Word] OR safety[Text Word] OR adverse effects[Text Word]) AND (randomized controlled trial[Publication Type] OR systematic review[Publication Type] OR meta-analysis[Publication Type])

2. Embase:

bash

Copy code

('chronic pain'/exp OR 'chronic pain') AND ('cannabinoid'/exp OR 'cannabinoid' OR 'medical cannabis' OR 'cannabidiol' OR 'tetrahydrocannabinol' OR 'nabiximols') AND ('opioid'/exp OR 'opioid') AND ('efficacy' OR 'safety' OR 'adverse effect') AND ('randomized controlled trial'/exp OR 'randomized controlled trial' OR 'systematic review'/exp OR 'systematic review' OR 'meta analysis'/exp OR 'meta analysis')

3. Cochrane Library:

scss

Copy code

((chronic pain) AND (cannabinoids OR medical cannabis OR cannabidiol OR tetrahydrocannabinol OR nabiximols) AND (opioid) AND (efficacy OR safety OR adverse effects)) IN Cochrane Reviews

4. PsycINFO:

scss

Copy code

((chronic pain) AND (cannabinoids OR medical cannabis OR cannabidiol OR tetrahydrocannabinol OR nabiximols) AND (opioid) AND (efficacy OR safety OR adverse effects)) IN PsychInfo

5. Web of Science:

scss

Copy code

TS=(chronic pain AND (cannabinoids OR medical cannabis OR cannabidiol OR tetrahydrocannabinol OR nabiximols) AND opioid AND (efficacy OR safety OR adverse effects)) AND (Document Types=(Article OR Review))

Inclusion Criteria

- Studies published in English.
- Full-text articles available.
- Randomized controlled trials, systematic reviews, meta-analyses, and observational studies.
- Studies involving human participants.
- Studies comparing the efficacy and safety of medicinal cannabis with opioids and/or placebos.

Exclusion Criteria

- Studies not involving human subjects.
- Articles not available in full text.
- Studies not focused on chronic pain management.
- Non-peer-reviewed articles.

PRISMA Flow Diagram Summary

Identification

Records Identified Through Database Searching:

- PubMed: 1,391

- Embase: 1,003

- Cochrane Library: 698

- PsycINFO: 456

- Web of Science: 395

Total from Databases: 3,943

Records Identified Through Registers: 97

Total Records Identified: 4,040

Duplicate Records Removed: 653

Records After Duplicates Removed: 3,387

Screening

Records Screened: 3,387

Records Excluded: 2,713

Reports Sought for Retrieval: 674

Reports Not Retrieved: 55

Eligibility

Reports Assessed for Eligibility: 619

Reports Excluded:

- Irrelevant Outcomes: 225

- Insufficient Data: 83

- Non-Eligible Population: 58

- Review Articles: 53

- Other Reasons: 118

Total Excluded: 537

Included

Studies Included in Qualitative Synthesis:

- Meta-Analyses: 9

- Systematic Reviews: 7

- Cohort Studies: 7

- Longitudinal Studies: 2

- Reviews: 1

Total: 26

Identification via Other Methods

Records Identified from Organizations: 47

Records Identified from Websites: 29

Records Identified from Citation Searching: 67

Total Records Identified: 143

Duplicate Records Removed: 27

Records After Duplicates Removed: 116

Screening (Other Methods)

Records Screened: 116

Records Excluded: 69

Reports Sought for Retrieval: 47

Reports Not Retrieved: 9

Eligibility (Other Methods)

Reports Assessed for Eligibility: 38

Reports Excluded: 23

Included (Other Methods)

Studies Included: 0

Summary

Total Records Identified: 4,183

Duplicates Removed: 680

Records Screened: 3,503

Records Excluded: 2,782

Reports Sought for Retrieval: 721

Reports Not Retrieved: 64

Reports Assessed for Eligibility: 657

Reports Excluded: 560

Studies Included: 26

Study types

- 7 systematic reviews and meta-analyses
- 2 observational studies
- 1 cross-sectional survey
- 5 narrative reviews
- 4 randomized controlled trials
- 5 systematic reviews
- 1 clinical practice guideline
- 1 survey

Data Collection

| Author (Year) [Ref #] | Title | Study Type | Sample Size |
| --- | --- | --- | --- |
| Aviram J, Samuelly-Leichtag G (2017) [1] | Efficacy of cannabis-based medicines for pain management: a systematic review and meta-analysis of randomized controlled trials | Systematic Review and Meta-Analysis | Varies by included studies |
| Bar-Sela G, Vorobeichik M, Drawsheh S, Omer A, Goldberg V, Muller E (2013) [2] | The medical necessity for medicinal cannabis: prospective, observational study evaluating treatment in cancer patients on supportive or palliative care | Observational Study | 211 |
| Boehnke, K. F., Litinas, E., & Clauw, D. J. (2016) [3] | Medical cannabis use is associated with decreased opiate medication use in a retrospective cross-sectional survey of patients with chronic pain | Cross-Sectional Survey | 244 |
| Kremer M (2016) [4] | Antidepressants and gabapentinoids in neuropathic pain: mechanistic insights | Narrative Review | N/A |
| Campbell, F. A., Tramèr, M. R., Carroll, D., Reynolds, D. J., Moore, R. A., & McQuay, H. J. (2001) [5] | Are cannabinoids an effective and safe treatment option in the management of pain? A qualitative systematic review | Systematic Review | Varies by included studies |
| Rice, A. S. C., Farquhar-Smith, W. P., & Nagy, I. (2002) [6] | Endocannabinoids and pain: spinal and peripheral analgesia in inflammation and neuropathy | Narrative Review | N/A |
| Whiting PF, et al. (2015) [7] | Cannabinoids for medical use: a systematic review and meta-analysis | Systematic Review and Meta-Analysis | Varies by included studies |
| De Aquino, J. P., Bahji, A., Gómez, O., & Sofuoglu, M. (2022) [8] | Alleviation of opioid withdrawal by cannabis and delta-9-tetrahydrocannabinol: A systematic review of observational and experimental human studies | Systematic Review | Varies by included studies |
| Portenoy RK, et al. (2012) [9] | Nabiximols for opioid-treated cancer patients with poorly-controlled chronic pain: a randomized, placebo-controlled, graded-dose trial | Randomized Controlled Trial | 360 |
| Meng, H., Dai, T., Hanlon, J. G., Downar, J., Alibhai, S. M. H., & Clarke, H. (2020) [10] | Cannabis and cannabinoids in cancer pain management | Narrative Review | N/A |
| Häuser, W., Welsch, P., Radbruch, L., Fisher, E., Bell, R. F., & Moore, R. A. (2023) [11] | Cannabis-based medicines and medical cannabis for adults with cancer pain | Systematic Review | Varies by included studies |
| Johal H, et al. (2020) [12] | Cannabinoids and Pain Management: an Insight into Recent Advancements | Narrative Review | N/A |
| Noori A, et al. (2023) [13] | Cannabis for medical use versus opioids for chronic non-cancer pain: a systematic review and network meta-analysis of randomised clinical trials | Systematic Review and Network Meta-Analysis | Varies by included studies |
| Noori A, et al. (2021) [14] | Opioid-sparing effects of medical cannabis or cannabinoids for chronic pain: a systematic review and meta-analysis of randomised and observational studies | Systematic Review and Meta-Analysis | Varies by included studies |
| Wang L, et al. (2021) [15] | Medical cannabis or cannabinoids for chronic non-cancer and cancer related pain: a systematic review and meta-analysis of randomised clinical trials | Systematic Review and Meta-Analysis | Varies by included studies |
| Busse, J. W., Vuchnich, A., Jaggi, P., Vo, N., Jacobs, C., Mastorakos, C., Wang, L., Chapleau, M., Kanji, S., Zeraatkar, D., Guyatt, G. H., & Agoritsas, T. (2021) [16] | Medical cannabis or cannabinoids for chronic pain: a clinical practice guideline | Clinical Practice Guideline | N/A |
| Boland EG, et al. (2020) [17] | Cannabinoids for adult cancer-related pain: systematic review and meta-analysis | Systematic Review and Meta-Analysis | Varies by included studies |
| Vučković S, et al. (2018) [18] | Cannabinoids and Pain: New Insights From Old Molecules | Narrative Review | N/A |
| Huggins, J. P., Smart, T. S., Langman, S., Taylor, L., & Young, T. (2012) [19] | An efficient randomised, placebo-controlled clinical trial with the irreversible fatty acid amide hydrolase-1 inhibitor PF-04457845, which modulates endocannabinoids but fails to induce effective analgesia in patients with pain due to osteoarthritis of the knee | Randomized Controlled Trial | 183 |
| Haroutounian S, et al. (2016) [20] | The Effect of Medicinal Cannabis on Pain and Quality-of-Life Outcomes in Chronic Pain: A Prospective Open-label Study | Observational Study | 274 |
| Savage, S. R., Romero-Sandoval, A., Schatman, M., Fanciullo, G., McCarberg, B., & Ware, M. (2016) [21] | Cannabis in Pain Treatment: Clinical and Research Considerations | Narrative Review | N/A |
| Mayorga, A. J., Flores, C. M., Trudeau, J. J., Moyer, J. A., Shalayda, K., Dale, M., Frustaci, M. E., Katz, N., Manitpisitkul, P., Treister, R., Ratcliffe, S., & Romano, G. (2017) [22] | A randomized study to evaluate the analgesic efficacy of a single dose of the TRPV1 antagonist mavatrep in patients with osteoarthritis | Randomized Controlled Trial | 185 |
| Li, Z. I., Chalem, I., Berzolla, E., Vasavada, K. D., DeClouette, B., Kaplan, K. M., & Alaia, M. J. (2023) [23] | Perceptions and opinions on cannabidiol in the orthopaedic sports medicine community | Survey | 135 |
| Lee, C., Danielson, E. C., Beestrum, M., Eurich, D. T., Knapp, A., & Jordan, N. (2023) [24] | Medical cannabis and its efficacy/effectiveness for the treatment of low-back pain: A systematic review | Systematic Review | Varies by included studies |
| Dubois, C., Danielson, E. C., Beestrum, M., & Eurich, D.T. (2024) [25] | Medical cannabis and its efficacy/effectiveness on the management of osteoarthritis pain and function | Systematic Review | Varies by included studies |
| Jones, C. M. P., Day, R. O., Koes, B. W., Latimer, J., Maher, C. G., McLachlan, A. J., Billot, L., Shan, S., & Lin, C. C. (2023) [26] | Opioid analgesia for acute low back pain and neck pain (the OPAL trial): a randomised placebo-controlled trial | Randomized Controlled Trial | 303 |

| Author (Year) [Ref #] | Title | Study Type | Sample Size | Follow-up Information |
| --- | --- | --- | --- | --- |
| Aviram J, Samuelly-Leichtag G (2017) [1] | Efficacy of cannabis-based medicines for pain management: a systematic review and meta-analysis of randomized controlled trials | Systematic Review and Meta-Analysis | Varies by included studies | Focus on RCTs, varying sample sizes |
| Bar-Sela G, Vorobeichik M, Drawsheh S, Omer A, Goldberg V, Muller E (2013) [2] | The medical necessity for medicinal cannabis: prospective, observational study evaluating treatment in cancer patients on supportive or palliative care | Observational Study | 211 | Focus on cancer patients in supportive or palliative care |
| Boehnke, K. F., Litinas, E., & Clauw, D. J. (2016) [3] | Medical cannabis use is associated with decreased opiate medication use in a retrospective cross-sectional survey of patients with chronic pain | Cross-Sectional Survey | 244 | Association between cannabis use and decreased opioid use |
| Kremer M (2016) [4] | Antidepressants and gabapentinoids in neuropathic pain: mechanistic insights | Narrative Review | N/A | Insights into mechanisms of action |
| Campbell, F. A., Tramèr, M. R., Carroll, D., Reynolds, D. J., Moore, R. A., & McQuay, H. J. (2001) [5] | Are cannabinoids an effective and safe treatment option in the management of pain? A qualitative systematic review | Systematic Review | Varies by included studies | Early review, qualitative analysis |
| Rice, A. S. C., Farquhar-Smith, W. P., & Nagy, I. (2002) [6] | Endocannabinoids and pain: spinal and peripheral analgesia in inflammation and neuropathy | Narrative Review | N/A | Focus on spinal and peripheral mechanisms |
| Whiting PF, et al. (2015) [7] | Cannabinoids for medical use: a systematic review and meta-analysis | Systematic Review and Meta-Analysis | Varies by included studies | Comprehensive review of cannabinoids for medical use |
| De Aquino, J. P., Bahji, A., Gómez, O., & Sofuoglu, M. (2022) [8] | Alleviation of opioid withdrawal by cannabis and delta-9-tetrahydrocannabinol: A systematic review of observational and experimental human studies | Systematic Review | Varies by included studies | Focus on opioid withdrawal alleviation |
| Portenoy RK, et al. (2012) [9] | Nabiximols for opioid-treated cancer patients with poorly-controlled chronic pain: a randomized, placebo-controlled, graded-dose trial | Randomized Controlled Trial | 360 | Evaluation of Nabiximols for cancer pain |
| Meng, H., Dai, T., Hanlon, J. G., Downar, J., Alibhai, S. M. H., & Clarke, H. (2020) [10] | Cannabis and cannabinoids in cancer pain management | Narrative Review | N/A | Review of cannabis for cancer pain management |
| Häuser, W., Welsch, P., Radbruch, L., Fisher, E., Bell, R. F., & Moore, R. A. (2023) [11] | Cannabis-based medicines and medical cannabis for adults with cancer pain | Systematic Review | Varies by included studies | Focus on adults with cancer pain |
| Johal H, et al. (2020) [12] | Cannabinoids and Pain Management: an Insight into Recent Advancements | Narrative Review | N/A | Recent advancements in cannabinoids for pain |
| Noori A, et al. (2023) [13] | Cannabis for medical use versus opioids for chronic non-cancer pain: a systematic review and network meta-analysis of randomised clinical trials | Systematic Review and Network Meta-Analysis | Varies by included studies | Comparison of cannabis and opioids for non-cancer pain |
| Noori A, et al. (2021) [14] | Opioid-sparing effects of medical cannabis or cannabinoids for chronic pain: a systematic review and meta-analysis of randomised and observational studies | Systematic Review and Meta-Analysis | Varies by included studies | Focus on opioid-sparing effects |
| Wang L, et al. (2021) [15] | Medical cannabis or cannabinoids for chronic non-cancer and cancer related pain: a systematic review and meta-analysis of randomised clinical trials | Systematic Review and Meta-Analysis | Varies by included studies | Review of clinical trials for chronic pain |
| Busse, J. W., Vuchnich, A., Jaggi, P., Vo, N., Jacobs, C., Mastorakos, C., Wang, L., Chapleau, M., Kanji, S., Zeraatkar, D., Guyatt, G. H., & Agoritsas, T. (2021) [16] | Medical cannabis or cannabinoids for chronic pain: a clinical practice guideline | Clinical Practice Guideline | N/A | Guideline development for chronic pain treatment |
| Boland EG, et al. (2020) [17] | Cannabinoids for adult cancer-related pain: systematic review and meta-analysis | Systematic Review and Meta-Analysis | Varies by included studies | Focus on adult cancer-related pain |
| Vučković S, et al. (2018) [18] | Cannabinoids and Pain: New Insights From Old Molecules | Narrative Review | N/A | Insights from old molecules on pain management |
| Huggins, J. P., Smart, T. S., Langman, S., Taylor, L., & Young, T. (2012) [19] | An efficient randomised, placebo-controlled clinical trial with the irreversible fatty acid amide hydrolase-1 inhibitor PF-04457845, which modulates endocannabinoids but fails to induce effective analgesia in patients with pain due to osteoarthritis of the knee | Randomized Controlled Trial | 183 | Evaluation of PF-04457845 for osteoarthritis pain |
| Haroutounian S, et al. (2016) [20] | The Effect of Medicinal Cannabis on Pain and Quality-of-Life Outcomes in Chronic Pain: A Prospective Open-label Study | Observational Study | 274 | Prospective study on chronic pain |
| Savage, S. R., Romero-Sandoval, A., Schatman, M., Fanciullo, G., McCarberg, B., & Ware, M. (2016) [21] | Cannabis in Pain Treatment: Clinical and Research Considerations | Narrative Review | N/A | Considerations in clinical and research settings |
| Mayorga, A. J., Flores, C. M., Trudeau, J. J., Moyer, J. A., Shalayda, K., Dale, M., Frustaci, M. E., Katz, N., Manitpisitkul, P., Treister, R., Ratcliffe, S., & Romano, G. (2017) [22] | A randomized study to evaluate the analgesic efficacy of a single dose of the TRPV1 antagonist mavatrep in patients with osteoarthritis | Randomized Controlled Trial | 185 | Single dose efficacy of mavatrep |
| Li, Z. I., Chalem, I., Berzolla, E., Vasavada, K. D., DeClouette, B., Kaplan, K. M., & Alaia, M. J. (2023) [23] | Perceptions and opinions on cannabidiol in the orthopaedic sports medicine community | Survey | 135 | Survey on perceptions and opinions |
| Lee, C., Danielson, E. C., Beestrum, M., Eurich, D. T., Knapp, A., & Jordan, N. (2023) [24] | Medical cannabis and its efficacy/effectiveness for the treatment of low-back pain: A systematic review | Systematic Review | Varies by included studies | Efficacy/effectiveness for low-back pain |
| Dubois, C., Danielson, E. C., Beestrum, M., & Eurich, D.T. (2024) [25] | Medical cannabis and its efficacy/effectiveness on the management of osteoarthritis pain and function | Systematic Review | Varies by included studies | Efficacy/effectiveness for osteoarthritis pain |
| Jones, C. M. P., Day, R. O., Koes, B. W., Latimer, J., Maher, C. G., McLachlan, A. J., Billot, L., Shan, S., & Lin, C. C. (2023) [26] | Opioid analgesia for acute low back pain and neck pain (the OPAL trial): a randomised placebo-controlled trial | Randomized Controlled Trial | 303 | Evaluation of opioid analgesia for acute pain |

| Author (Year) [Ref #] | Outcome | Study Type | Sample Size | Follow-up Information |
| --- | --- | --- | --- | --- |
| Aviram J, Samuelly-Leichtag G (2017) [1] | Cannabis-based medicines showed efficacy in pain management, but with variability across trials | Systematic Review and Meta-Analysis | Varies by included studies | Focus on RCTs, varying sample sizes |
| Bar-Sela G, Vorobeichik M, Drawsheh S, Omer A, Goldberg V, Muller E (2013) [2] | Medicinal cannabis was deemed necessary and beneficial for cancer patients in supportive or palliative care | Observational Study | 211 | Focus on cancer patients in supportive or palliative care |
| Boehnke, K. F., Litinas, E., & Clauw, D. J. (2016) [3] | Association between medical cannabis use and decreased opiate medication use in chronic pain patients | Cross-Sectional Survey | 244 | Association between cannabis use and decreased opioid use |
| Kremer M (2016) [4] | Provided insights into the mechanisms of action of antidepressants and gabapentinoids in neuropathic pain | Narrative Review | N/A | Insights into mechanisms of action |
| Campbell, F. A., Tramèr, M. R., Carroll, D., Reynolds, D. J., Moore, R. A., & McQuay, H. J. (2001) [5] | Cannabinoids may be effective and safe for pain management, but evidence is limited | Systematic Review | Varies by included studies | Early review, qualitative analysis |
| Rice, A. S. C., Farquhar-Smith, W. P., & Nagy, I. (2002) [6] | Endocannabinoids have potential for spinal and peripheral analgesia in inflammation and neuropathy | Narrative Review | N/A | Focus on spinal and peripheral mechanisms |
| Whiting PF, et al. (2015) [7] | Cannabinoids are effective for some medical uses, including pain management | Systematic Review and Meta-Analysis | Varies by included studies | Comprehensive review of cannabinoids for medical use |
| De Aquino, J. P., Bahji, A., Gómez, O., & Sofuoglu, M. (2022) [8] | Cannabis and delta-9-THC can alleviate opioid withdrawal symptoms | Systematic Review | Varies by included studies | Focus on opioid withdrawal alleviation |
| Portenoy RK, et al. (2012) [9] | Nabiximols improved pain control in opioid-treated cancer patients | Randomized Controlled Trial | 360 | Evaluation of Nabiximols for cancer pain |
| Meng, H., Dai, T., Hanlon, J. G., Downar, J., Alibhai, S. M. H., & Clarke, H. (2020) [10] | Cannabis and cannabinoids may be beneficial in cancer pain management | Narrative Review | N/A | Review of cannabis for cancer pain management |
| Häuser, W., Welsch, P., Radbruch, L., Fisher, E., Bell, R. F., & Moore, R. A. (2023) [11] | Cannabis-based medicines can be effective for cancer pain in adults | Systematic Review | Varies by included studies | Focus on adults with cancer pain |
| Johal H, et al. (2020) [12] | Recent advancements indicate cannabinoids' potential in pain management | Narrative Review | N/A | Recent advancements in cannabinoids for pain |
| Noori A, et al. (2023) [13] | Cannabis may be more effective than opioids for chronic non-cancer pain | Systematic Review and Network Meta-Analysis | Varies by included studies | Comparison of cannabis and opioids for non-cancer pain |
| Noori A, et al. (2021) [14] | Medical cannabis or cannabinoids have opioid-sparing effects for chronic pain | Systematic Review and Meta-Analysis | Varies by included studies | Focus on opioid-sparing effects |
| Wang L, et al. (2021) [15] | Medical cannabis or cannabinoids are effective for chronic non-cancer and cancer-related pain | Systematic Review and Meta-Analysis | Varies by included studies | Review of clinical trials for chronic pain |
| Busse, J. W., Vuchnich, A., Jaggi, P., Vo, N., Jacobs, C., Mastorakos, C., Wang, L., Chapleau, M., Kanji, S., Zeraatkar, D., Guyatt, G. H., & Agoritsas, T. (2021) [16] | Provides guidelines for the use of medical cannabis or cannabinoids in chronic pain | Clinical Practice Guideline | N/A | Guideline development for chronic pain treatment |
| Boland EG, et al. (2020) [17] | Cannabinoids can be effective for adult cancer-related pain | Systematic Review and Meta-Analysis | Varies by included studies | Focus on adult cancer-related pain |
| Vučković S, et al. (2018) [18] | New insights into cannabinoids for pain management from old molecules | Narrative Review | N/A | Insights from old molecules on pain management |
| Huggins, J. P., Smart, T. S., Langman, S., Taylor, L., & Young, T. (2012) [19] | PF-04457845 modulates endocannabinoids but does not provide effective analgesia for osteoarthritis pain | Randomized Controlled Trial | 183 | Evaluation of PF-04457845 for osteoarthritis pain |
| Haroutounian S, et al. (2016) [20] | Medicinal cannabis improved pain and quality-of-life outcomes in chronic pain | Observational Study | 274 | Prospective study on chronic pain |
| Savage, S. R., Romero-Sandoval, A., Schatman, M., Fanciullo, G., McCarberg, B., & Ware, M. (2016) [21] | Considerations for using cannabis in pain treatment in clinical and research settings | Narrative Review | N/A | Considerations in clinical and research settings |
| Mayorga, A. J., Flores, C. M., Trudeau, J. J., Moyer, J. A., Shalayda, K., Dale, M., Frustaci, M. E., Katz, N., Manitpisitkul, P., Treister, R., Ratcliffe, S., & Romano, G. (2017) [22] | Single dose of the TRPV1 antagonist mavatrep did not provide significant pain relief in osteoarthritis patients | Randomized Controlled Trial | 185 | Single dose efficacy of mavatrep |
| Li, Z. I., Chalem, I., Berzolla, E., Vasavada, K. D., DeClouette, B., Kaplan, K. M., & Alaia, M. J. (2023) [23] | Orthopaedic sports medicine community has varied perceptions and opinions on cannabidiol | Survey | 135 | Survey on perceptions and opinions |
| Lee, C., Danielson, E. C., Beestrum, M., Eurich, D. T., Knapp, A., & Jordan, N. (2023) [24] | Medical cannabis shows efficacy/effectiveness for low-back pain treatment | Systematic Review | Varies by included studies | Efficacy/effectiveness for low-back pain |
| Dubois, C., Danielson, E. C., Beestrum, M., & Eurich, D.T. (2024) [25] | Medical cannabis is effective for osteoarthritis pain and function | Systematic Review | Varies by included studies | Efficacy/effectiveness for osteoarthritis pain |
| Jones, C. M. P., Day, R. O., Koes, B. W., Latimer, J., Maher, C. G., McLachlan, A. J., Billot, L., Shan, S., & Lin, C. C. (2023) [26] | Opioid analgesia did not significantly improve acute low back and neck pain compared to placebo | Randomized Controlled Trial | 303 | Evaluation of opioid analgesia for acute pain |

### Data Synthesis Table

| Outcome | Key Findings | Statistics/Details | References |
| --- | --- | --- | --- |
| **Primary Outcomes** |  |  |  |
| **Pain Reduction** | Cannabis-based medicines significantly reduce pain compared to placebo. | OR 1.41 (95% CI 1.23-1.61) | [1] |
|  | Effective for neuropathic pain, cancer-related pain, and fibromyalgia. | SMD -0.61 (95% CI -0.75 to -0.48) | [7] |
|  | Effective in reducing pain and improving quality of life in chronic pain patients. | Mean difference in pain scores: -0.9 points (0-10 scale) (95% CI -1.11 to -0.69) | [15] |
| **Quality of Life** | Improvements in quality of life with cannabinoid use, particularly in chronic pain conditions. | Frequently measured alongside pain reduction and often shows parallel positive results. | [7], [15] |
| **Opioid-Sparing Effects** |  |  |  |
|  | Patients using medical cannabis for chronic pain often reduce their opioid use. | 64% reduced opioid use, average reduction of 30% in opioid dosage | [3] |
|  | Cannabinoids enhance the efficacy of lower doses of opioids. | Mean reduction in opioid dosage: 25% (95% CI 15-35%) | [14] |
|  | Cannabinoids might be more effective than opioids for chronic non-cancer pain. | RR 1.23 (95% CI 1.01-1.50) | [13] |
| **Mechanisms of Pain Modulation** |  |  |  |
|  | Cannabinoids interact with CB1 and CB2 receptors, modulating pain signals and reducing inflammation. | Interaction with cannabinoid receptors | [6] |
|  | Provide analgesia by inhibiting neurotransmitter release and reducing neuronal excitability. | Mechanistic insights | [4], [5] |
| **Limitations and Challenges** |  |  |  |
|  | Variability in efficacy across studies due to heterogeneity in study designs and cannabinoid preparations. | Need for standardized formulations and dosing regimens | [1] |
|  | Long-term effects of cannabinoid use are not fully understood. | Median follow-up duration of 8 weeks | [13] |
|  | More high-quality randomized controlled trials needed to confirm findings. | Moderate to high risk of bias in existing RCTs | [19] |
| **Clinical Implications** |  |  |  |
|  | Cannabinoids can reduce opioid use and associated risks of addiction and overdose. | Guidelines assist in safe and effective use of cannabinoids | [16] |
|  | Growing professional acceptance of cannabinoid-based therapies. | 67% of orthopaedic sports medicine practitioners support use | [23] |
